# Demographics of people who transmit HIV-1 in Zambia: a molecular epidemiology analysis in the HPTN-071 PopART study

**DOI:** 10.1101/2021.10.04.21263560

**Authors:** Matthew Hall, Tanya Golubchik, David Bonsall, Lucie Abeler-Dörner, Mohammed Limbada, Barry Kosloff, Ab Schaap, Mariateresa de Cesare, George MacIntyre-Cockett, Newton Otecko, William Probert, Oliver Ratmann, Ana Bulas Cruz, Estelle Piwowar-Manning, David N Burns, Myron S Cohen, Deborah J Donnell, Susan H Eshleman, Musonda Simwinga, Sarah Fidler, Richard Hayes, Helen Ayles, Christophe Fraser, the HPTN 071 (PopART) Phylogenetics protocol team and the PANGEA consortium

**Affiliations:** Big Data Institute and Pandemic Sciences Institute, Nuffield Department of Medicine, University of Oxford, Oxford, UK; Sydney Infectious Diseases Institute, School of Medical Sciences, Faculty of Medicine and Health, University of Sydney; Wellcome Centre for Human Genetics, Nuffield Department of Medicine, University of Oxford, Oxford, UK; Zambart, University of Zambia, Lusaka, Zambia; Department of Clinical Research, London School of Hygiene and Tropical Medicine, London, United Kingdom; Department of Mathematics, Imperial College, London, UK; Department of Pathology, Johns Hopkins University School of Medicine, Baltimore, Maryland 21205, USA; Division of AIDS, National Institute of Allergy and Infectious Diseases, National Institutes of Health, Rockville, Maryland, United States of America; Department of Medicine, University of North Carolina at Chapel Hill, Chapel Hill, USA; Fred Hutchinson Cancer Research Center, Seattle, WA, USA; Department of Infectious Disease Epidemiology, Imperial College, London, UK; Department of Infectious Disease Epidemiology, London School of Hygiene and Tropical Medicine, London, United Kingdom

## Abstract

**Background:** In the last decade, universally available antiretroviral therapy (ART) has led to greatly improved health and survival of people living with HIV in sub-Saharan Africa, and appears to have contributed to reduced rates of new infections. Individuals acting as sources of infection need to be characterised to design effective prevention strategies.

**Methods:** We used viral genomes to investigate the demographic characteristics of sources of HIV-1 infection. Between 2014 and 2018, the HPTN 071 PopART study was conducted to quantify the public health benefits of ART. Viral samples from 7,124 study participants in Zambia were deep-sequenced as part of HPTN 071-02 PopART Phylogenetics, an ancillary study. We identified 300 likely HIV-1 transmission pairs and investigated the source individuals in those pairs to better understand transmission in the general population.

**Findings:** After demographic weighting, 59.4% of transmissions were male to female, with 43·2% (95% CI: 36·8%-49·7%) of transmissions being from males aged 25-40. Overall, men transmitted 2.09-fold (2·06-2·29) more infections per capita than women, a ratio peaking, when stratified by source age, at 5.88 (2·78-15·8) in the 35-39 age group. 17·4% of sources (12·5%-22·4%) carried viruses resistant to first-line ART. 12·9% (8·5%-17·3%) of transmissions linked individuals from different communities in the trial.

**Interpretation:** HIV-1 transmission in the HPTN 071 study communities comes from a wide range of age and sex groups, and that there is no outsized contribution of importation or drug resistance mutations to new infections. Men aged 25-40, under-served by current treatment and prevention service, should be prioritised for HIV testing and ART.

**Funding:** National Institute of Allergy and Infectious Diseases, US President’s Emergency Plan for AIDS Relief, International Initiative for Impact Evaluation, Bill & Melinda Gates Foundation, National Institute on Drug Abuse, and National Institute of Mental Health

**Research in context panel:** *Evidence before this study:* We searched PubMed, with no date or language filters, to identify previous quantitative studies investigating the role of age, sex, mobility and drug resistance mutations (separately or in combination) as drivers of new HIV transmissions in heterosexual transmission in sub-Saharan Africa. Observational studies were considered along with those using phylogenetic or mathematical modelling methodologies. In observational studies, having an older partner or a migrant partner is frequently identified as a risk factor for HIV acquisition, particularly in women, but this is a slightly separate question to the quantification of the overall frequency of these demographics amongst the sources of new infections. The most recent studies of drug resistance have shown an increasing prevalence, particularly in NNRTI resistance. The ability to use phylogenetics to investigate HIV transmission is reasonably recent, and the ability to use it to reconstruct who infected whom in transmission pairs is more recent still. One previous, influential study in South Africa posited a key role of men in their 30s in infecting very young women, and being themselves infected by women in their 30s, but that work did not use a methodology that was able to conclusively reconstruct direction of transmission. A more recent study in Botswana found similar age distributions for male and female sources of transmission, with the average individual being in their late 30s or early 40s. However, these ages were recorded at the time of study enrolment and do not take into account the time from infection to sampling. The paper also showed that, in that setting, the majority of transmissions were between members of the same community. A number of other phylogenetic studies have been concerned with the very specific dynamics of HIV-1 in the fishing communities of Lake Victoria. No previous phylogenetics work to our knowledge has considered the distribution of drug resistance mutations in sources, and none considered the interaction between source characteristics. We were unable to find any previous mathematical modelling studies for which the characterisation of sources according to these variables was a major focus.

*Added value of this study:* Our methodology uses a phylogenetic approach to identify likely transmission pairs and the direction of transmission between their members. We find a total of 300 pairs, larger than any previous study. We demonstrate a novel and simple new approach to accounting for potential sampling bias. We also employ a methodology that allows us to estimate the ages of the individuals involved at the time of transmission, rather than that of sampling, countering a key bias in previous approaches. Partly for this reason, our age profiles for sources peak at earlier ages than in previous work, with an average in the early 30s for male sources and mid-20s for females. We fail to confirm the existence of a “renewal cycle” of transmission involving a major contribution from women in older age groups. We also examine the contribution of outside-community transmission and drug resistant mutations, and, for the first time, show that these three characteristics (age/sex, migration, and drug resistance) operate on separate axes and do not cluster together. We use our results to calculate the relative contribution of male to female sources to transmission in age bands, finding that this grows to a peak in the 35-39 age group in which men are responsible for almost six times as many new infections per capita as women.

*Implications of all the available evidence:* The heterosexual HIV epidemic in sub-Saharan Africa appears to be maintained by transmissions from young women and slightly, but not dramatically, older men. The contributions of sources transmitting drug-resistant virus, or sources who reside outside a focal community, is not particularly large, and there is no disproportionate contribution from individuals who share any combination of a high-risk age group, a residence outside the community, and drug-resistant virus. In generalised HIV epidemics, it is tempting to attempt to identify particular demographic groups, of relatively small sizes, for whom intensive targeting of prevention measures will have a major effect on transmission in the general population. The current state of the evidence suggests that this may not be possible, as the demographic profile of sources of transmission is not dissimilar to that of the general population. While it may be more difficult and resource-intensive to design universal interventions for the whole population, there may be no shortcuts.

## Introduction

The last decade has seen a global transformation in HIV care, with the near universal availability of affordable and effective combination antiretroviral therapy (ART), which durably suppresses viral replication, prevents AIDS, and stops onward transmission of the virus. The discovery that combination ART blocks transmission led to the idea that an effective form of HIV prevention is HIV testing, followed by initiation of ART for infected individuals^1^.

The HPTN 071 (PopART) cluster-randomised trial^2^ evaluated whether a prevention package including universal testing and treatment would reduce HIV incidence in communities in Zambia and South Africa. The trial reported a 20% reduction of incidence in intervention arms. A further rapid and sustained decrease in new cases is needed to reach the ambitious UNAIDS goals for reduced incidence^3^.

The HPTN 071-02 (PopART) Phylogenetics Study was set up in the Zambian communities of HPTN 071. It aimed to use viral genomes to characterise sources of ongoing transmission, assess the effectiveness of the intervention had it been rolled out nationwide, and identify the most promising policies for prevention in the future.

HIV phylogenies can reconstruct the demographic and spatial history of transmission. In population-level analyses, phylogenetic inference has been used to describe the origin and global spread of HIV^4^, characterise transmission dynamics in concentrated epidemics^5^, and, building on studies examining individual transmission events^6^, analyse transmission in HIV prevention trials^7^. In sub-Saharan African (SSA) settings, phylogenetics has been used to look at patterns of clustering^8^ and spatial spread^9,10^.

Recent methodological innovations in phylogenetics have made it possible, in large studies with high sampling density and after deidentification, to identify probable transmission pairs^11^. In a SSA context, this approach has been successfully used to disentangle patterns of HIV-1 transmission surrounding Ugandan fishing communities on Lake Victoria^9,10^. Here, we apply it to HPTN 071-02 data to investigate the generalised epidemic in Zambia. We characterised probable sources of transmission by sex and estimated age at transmission, as well as whether they resided in a different community from their recipient, and if their dominant viral strain was resistant to first-line ART.

## Methods

### Trial participants

HPTN 071-02 was conducted in nine of twelve Zambian PopART communities and included participants from the Population Cohort (PC; used to monitor incidence in the community) and additional individuals attending health care facilities (HCF). The PC enrolled one randomly selected resident aged 15-44 per randomly selected household, irrespective of HIV status, between 2014 and 2017, for a total of 48,301 participants. At the HCF, individuals were approached and subsequently enrolled if they consented and were HIV+ individuals aged 18 or over who started treatment (or restarted after an interruption) between 2016 and 2018. Details can be found in the trial protocol^12^. Participants gave written consent and the study was approved by the medical ethics committees of the University of Zambia, Stellenbosch University, the London School of Hygiene and Tropical Medicine, Imperial College London, and the University of Oxford.

### Genome sequencing

Sequences for viral RNA in each sample were obtained using veSEQ-HIV, a bait-capture based quantitative high-throughput whole-genome deep-sequencing method with a sensitivity of >5,000 RNA copies/ml for whole genomes^13^.

### Bioinformatic processing

Sequence reads were filtered to remove human and bacterial sequences using Kraken^14^, and assembled into contigs using SPAdes v3.10.1^15^. Contigs were aligned to a curated alignment of 165 representative HIV-1 genomes from the Los Alamos National Laboratory HIV database (http://www.hiv.lanl.gov/) to generate a consensus sequence. Individual HIV-1 reads were then mapped onto this consensus. Per-sample HIV-1 genomes were assembled with *shiver* v1.3^16^. This generated a custom consensus sequence for each sample; we kept only samples for which at least 50% of consensus positions had non-missing data. Reads were coordinate-translated by *shiver* to bring them into alignment with the HXB2 reference genome. 910 of 6,864 samples were sequenced more than once; for these we combined all reads into a single pool.

### Identification of transmission pairs

A cluster analysis using HIV-TRACE^17^ with an intentionally generous threshold of 0·04 was performed on the consensus sequences to subdivide the dataset and thus reduce computational time for subsequent phylogenetic reconstruction. Identification of probable transmission pairs was then conducted on the *shiver*-aligned reads using *phyloscanner*^11^ (see appendix p1 for full *phyloscanner* procedure and p6 for all software settings).

### Estimation of time since infection

HIV-phyloTSI^18^ was used to estimate the infection date of each individual from their within-host phylogenetic data and sampling date. For pairs, this allowed us to further estimate when the transmission event would have taken place if either partner was the source, and how old both partners were at that time.

### Reconstruction of direction of transmission

The likely direction of transmission for each pair was assessed using two independent methods, firstly the phylogenetic topology^11^, and secondly comparison of estimated infection dates (see appendix, p1). Where both methods assigned the same direction of transmission, or one assigned a direction and the other returned an indeterminate result, we took the pair forwards as one with a determined direction of transmission. We excluded pairs where the recipient’s estimated time of infection was outside the timeframe of the PopART trial (November 2013 to December 2018).

### Classification of drug resistance mutations

Drug resistance (DR) to first-line adult ART was predicted based on detection of mutations from the Stanford HIV Drug Resistance Database scoring system (HIVdb version 8.9.1)^19^ with the following cut-offs: 0-14 -wild type/susceptible, 15-29 -low-level resistance, 30 and above -high-level resistance. See appendix, p1, for further details.

### Characteristics of sources and demographic weighting

For each directed pair, we characterised the source partner by sex, age group, viral DR mutations, and whether they were registered in a different community from their recipient. Sampling bias is an important consideration in a source attribution study of this sort. We observed that the study could be viewed as a survey in which recipients of infection provided information about their sources.

Thus, methods for adjusting for unrepresentative sampling from survey methodology are appropriate, and we used iterative proportional fitting (“raking”) to weight the population of recipients to be demographically representative of the HIV-positive population of individuals in the trial, in regard to four variables: sex, birth cohort, community, and marital status. The target population distribution of the former three variables was estimated from the PopART-IBM individual-based model^20^ while the last was calculated from the marital status of HIV-infected individuals in the 2018 Zambian Demographic and Health Survey^21^. Proportions of sources with a particular characteristic (e.g. age group) were then calculated by using the sum of their recipients’ weights as numerator, instead of the simple number of pairs. 95% confidence intervals for weighted proportions were calculated using Lyapunov’s central limit theorem.

## Results

Sampling yielded 6,864 genomic sequences. We analysed 5,611 for which 50% coverage of the consensus genome was obtained. (Figure 1, Table 1, Table S1). The *phyloscanner* procedure identified a total of 801 pairs, of which 469 were opposite-sex, 242 female-to-female and 90 male-to-male. The same-sex pairs are likely to be mostly or entirely erroneously linked (see Discussion), and as our focus was anyway on the heterosexual epidemic, we analysed the opposite-sex set only.

**Table 1:**
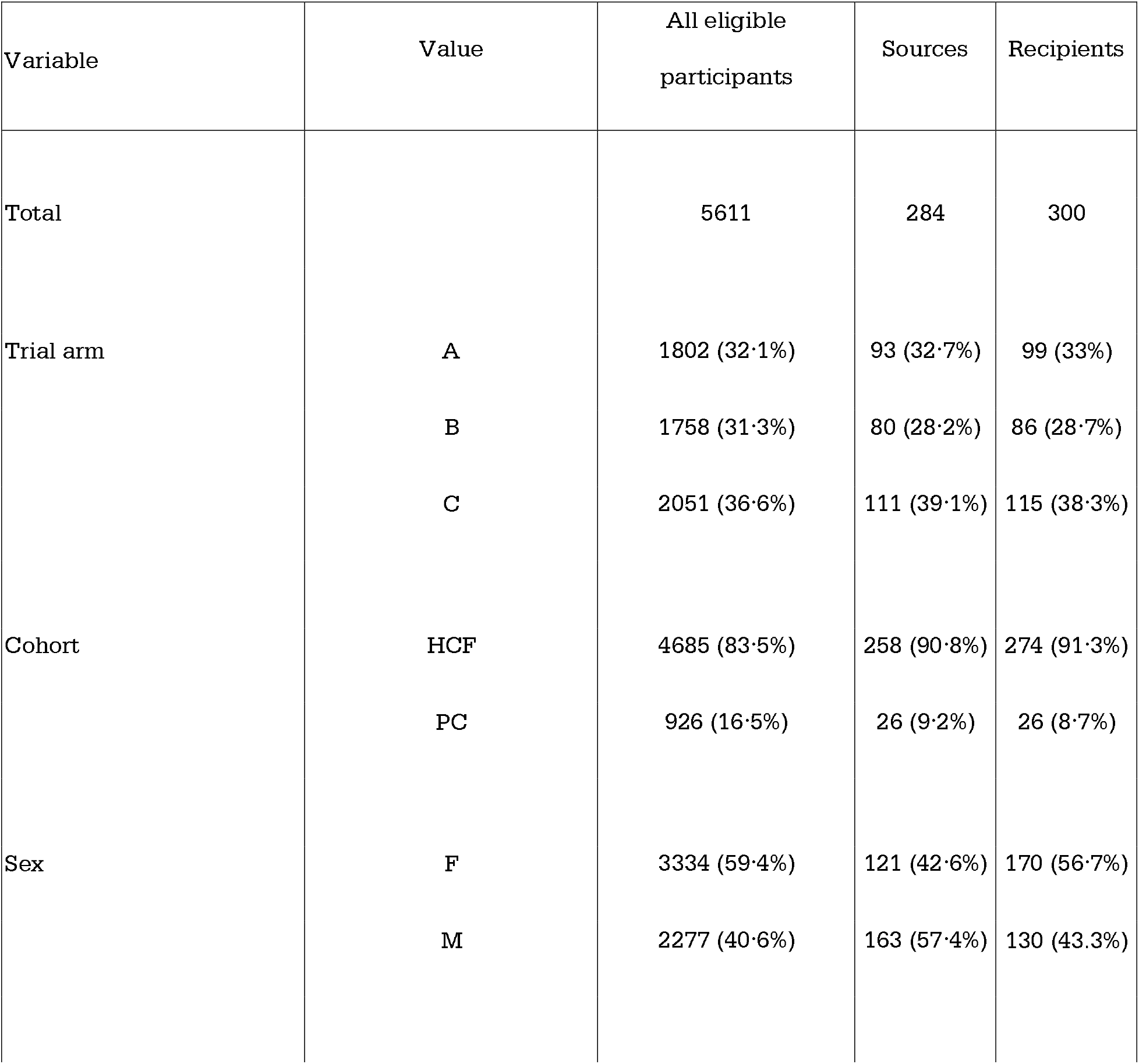
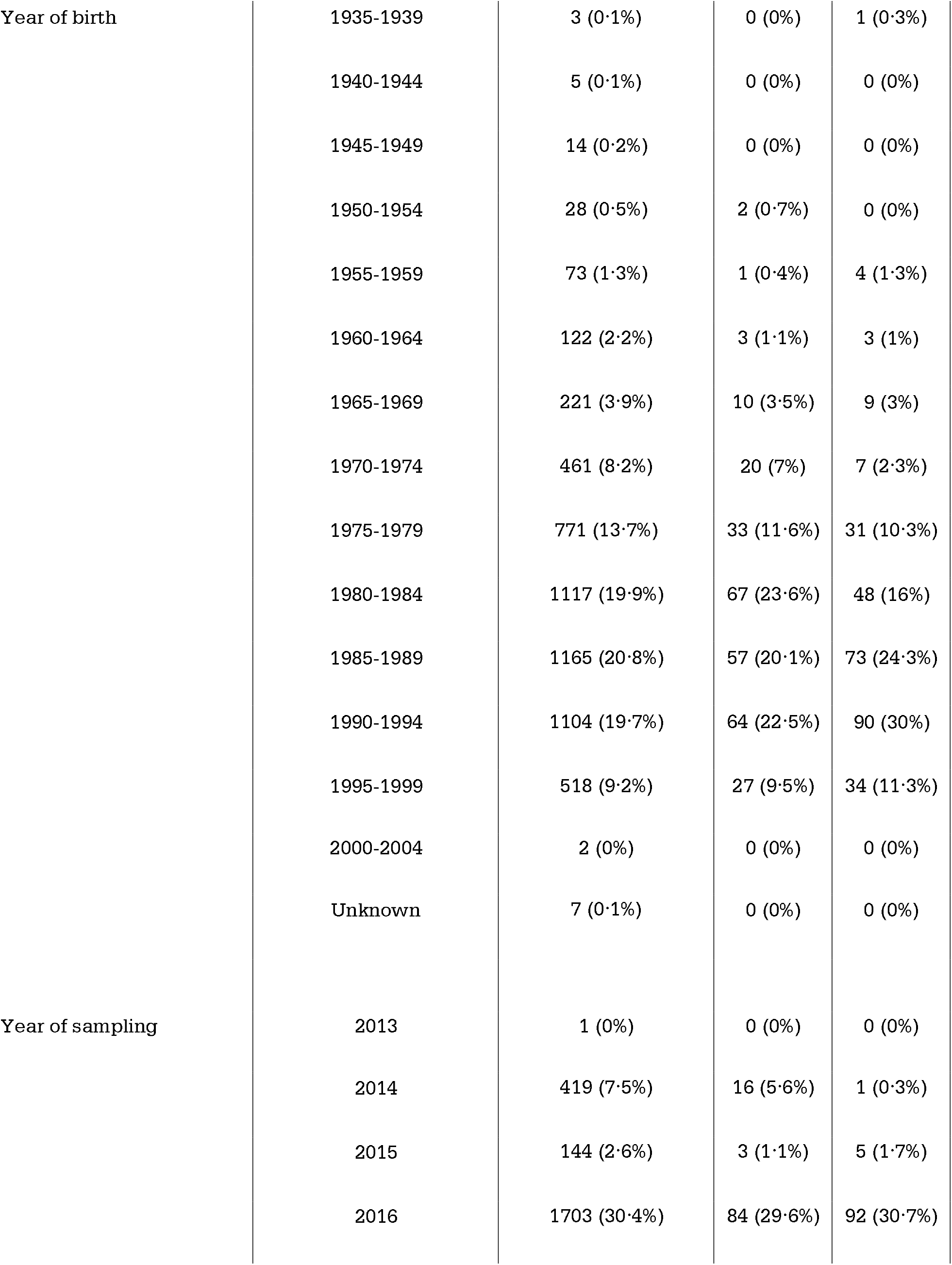
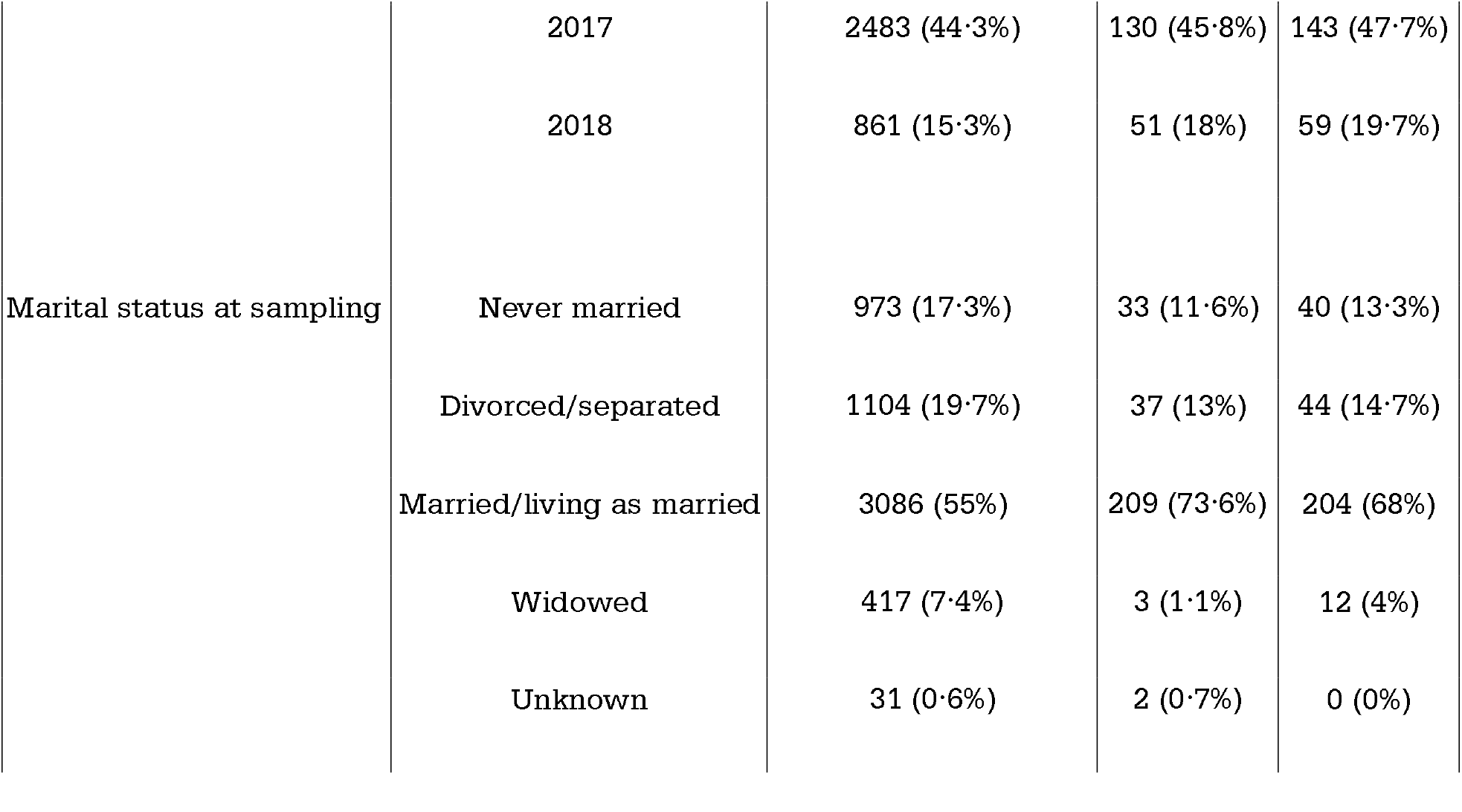
Demographic and other characteristics of all eligible participants, and probable sources and recipients in the reconstructed transmission pairs. 14 sources had multiple probable recipients.

**Fig. 1:**
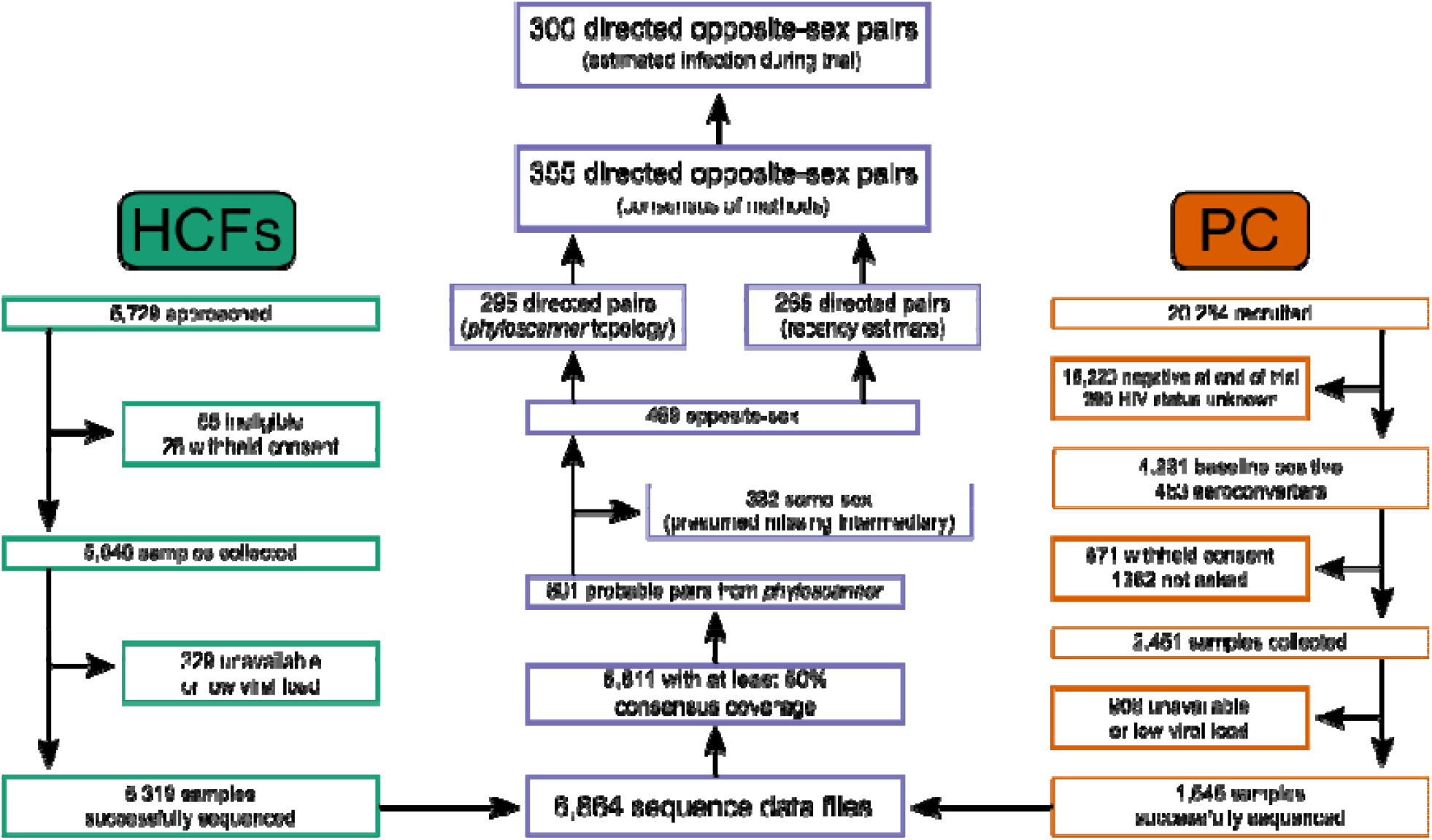
Flowchart depicting determination of transmission pairs from recruited participants from the 9 Zambian communities. “PC” refers to the population cohort used to evaluate the trial’s effect on HIV incidence, and “HCFs” to health care facilities in the trial communities.

The phylogenetic topology assigned a direction of transmission to 295 pairs, while 265 were called by comparison of the time of infection estimates. 145 were called by both methods, with 25 excluded due to conflicting results. When combined, the analysis yielded 355 probable directed transmission pairs. For 300, transmission was estimated to have occurred during the trial period (Figure 1). This was consistent with a power calculation conducted before the study to determine the number of transmission pairs required to characterise sources of infection (see appendix p1).

Table 1 outlines the characteristics of the individuals involved in the pairs. Note that there are fewer probable sources identified than recipients, as 14 individuals were reconstructed as infecting multiple others. We emphasise that the principal unit of our analysis is the transmission, and that characteristics such as age at time of transmission and the source residing in a different community to the recipient will not necessarily be consistent between two transmissions even with the same source. We are exploring the characteristics of the sources involved in 300 distinct transmissions, rather than those of 300 unique sources.

The age distributions of the individuals identified as probable sources, both male and female, showed the highest rates of transmission coming from individuals between 20 and 40 years of age: 72·7% (95% CI: 64·9%-80·6%) of male sources were between 25 and 40 at the estimated time of transmission, representing 43·2% (36·8%-49·7%) of all transmissions, while 71·5% (62·8%-80·2%) of female sources were between 20 and 35, representing 29% (23·1%-34·9%) (Figure 2A). (Unless otherwise stated, proportions and medians given in this section are all weighted to adjust for sampling bias as described above.) As estimated by PopART-IBM, men aged 25 to 40 represented only 19.4% of all infected individuals in 2017, while women aged 20 to 35 represented 27.3%. The median age at time of transmission was 32 years (range 20-64) for male sources and 25 (17-49) for female sources (Figure 2A). There was no evidence of a difference in the ages of sources (weighted ANOVA *p*=0·822 for male-to-female pairs, *p*=0·377 for female-to-male pairs) between trial arms.

**Figure 2:**
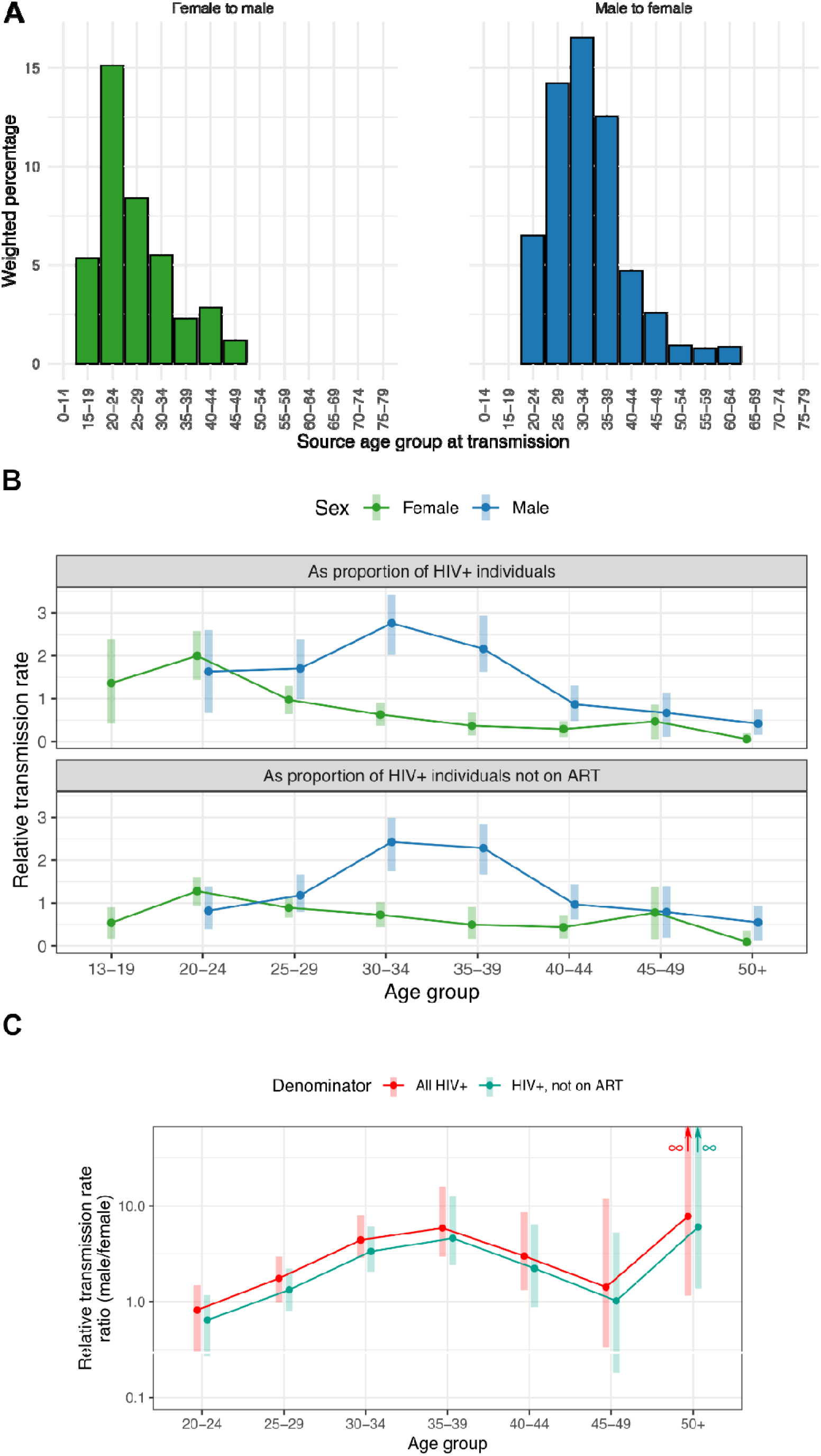
Age and sex characteristics of sources of transmission. (A) Age profile (at the estimated time of transmission) of the sources of infection for participants estimated to have been infected during the trial period. Counts were weighted to make the recipient population representative of the HIV positive population during the trial. (B) For different age and sex demographics, the ratio of the proportion of sources in that group to the proportion in the same group of (top) all HIV+ individuals or (bottom) all HIV+ individuals not on ART. Ages are calculated as of July 2017. Shaded bars are 95% confidence intervals determined by non-parametric bootstrap. We identified no male sources in the 13-19 age group and thus this estimate is omitted. (C) Ratios of the relative contributions of male and female sources by age group (as calculated above). Shaded bars are 95% confidence intervals as before. The bootstrapped confidence intervals for the 50+ age group extend upwards to infinity.

Pre-weighting, 56·7% of transmissions were from male to female participants; weighting brought this to 59·4%. We calculated relative transmission rates amongst demographic groups, defined as the ratio of the proportion of sources identified from a group to that group’s proportion of the overall HIV-positive population, as estimated by PopART-IBM (see appendix p2). For men, this was 1·44 (bootstrapped 95% CI: 1·43-1·5) while for women it was 0·69 (0·65-0·7). The ratio of these two numbers is 2·09 (2·06-2·29), indicating that HIV-positive males were contributing more than twice as many new infections as HIV-positive females, per capita. The ratio was 1·79 (1·78-1·95) when the group’s proportion of the HIV-positive population not on ART, instead of the total HIV-positive population, was used as a denominator, indicating that even when adjusting for ART coverage, men infected more women per capita than vice versa.

Figures 2B and 2C show these statistics for both denominators stratified by age group. The peak relative rate amongst females was observed in the 20-24 group (2·00 compared to the HIV+ population, 95% CI 1·41-2·67) while for males it was the 30-34 group (2·76; 2·09-3·37). The male-to-female ratio was 4·4 (2·8-8·25) in the 30-34 year age group and 5·88 (2·78-15·8) in the 35-39 year age group. This ratio was above 1 in all age groups other than 20-24 but the confidence interval included 1 for the 20-24, 25-29, and 45-49 groups for both denominators.

After omitting eleven individuals (3·8%) whose viruses had an unknown drug resistance profile (due to poor genome coverage at relevant sites), a large majority of sources (236/289) had drug-sensitive viruses, while 20 participants showed low-level resistance to first-line

ART and 33 HIV-1 showed high-level resistance (Figure 3A). The weighted proportions of individuals transmitting sensitive, low-level resistant and high-level resistant HIV-1 were 78·8% (73·5%-84.1), 6·75% (3·47%-10%) and 10·7% (6·63%-14·7%), respectively. Once again, there was no evidence of a difference in the distribution of mutations with respect to the trial arm of the recipient (chi-squared test *p*=0·321). For comparison, 79·7%, 11% and 9·3% of recipients had virus that was sensitive, low-level, or high-level resistant, in line with a previously published study on resistance^22^.

**Figure 3:**
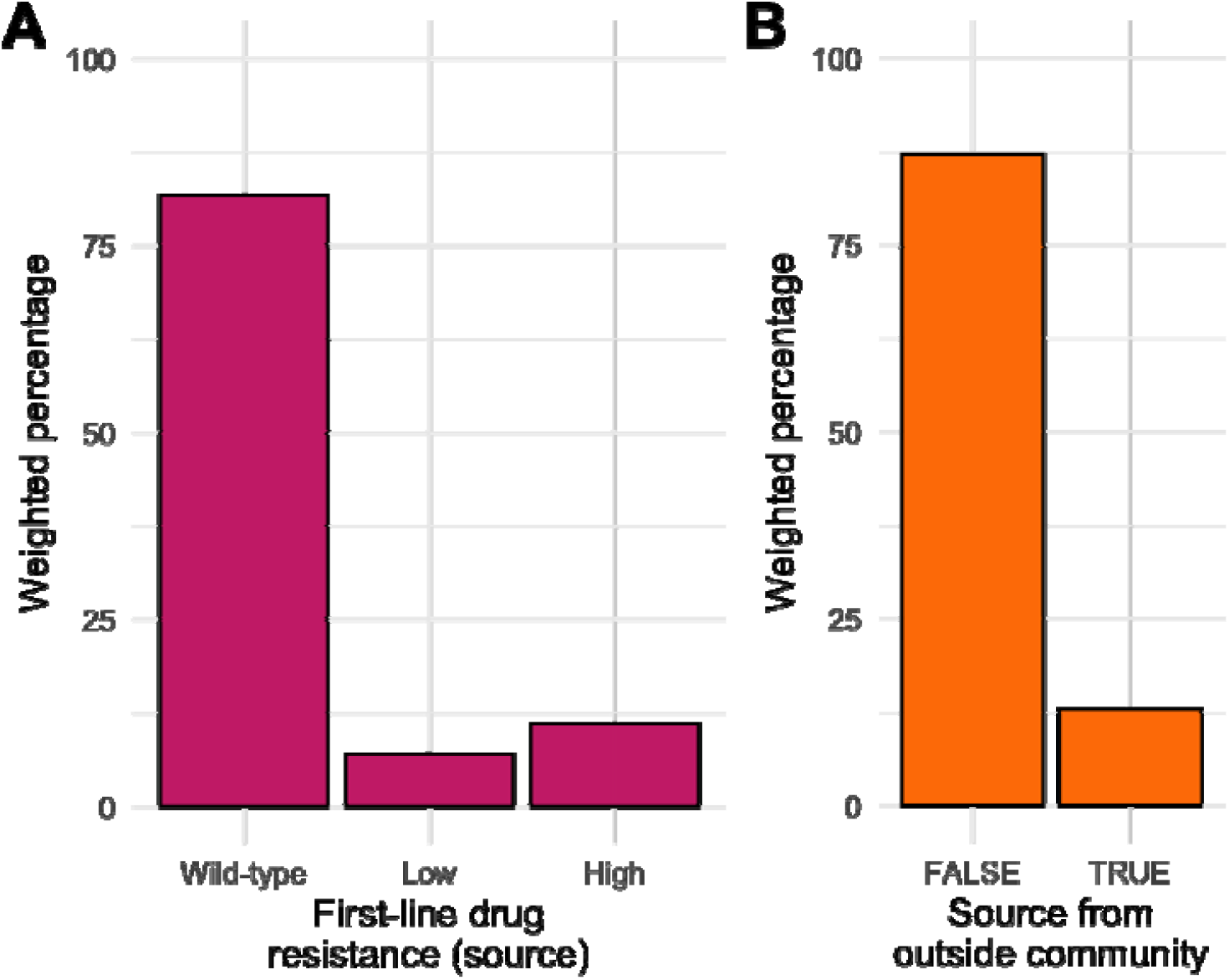
Other characteristics of source individuals. Counts were weighted to make the recipient population representative of the HIV positive population during the trial. (A) First-line drug resistance profiles. (B) Infections from sources residing in a different community from the recipient.

Lastly, we determined the fraction of transmission pairs for which the source lived in a different community from the recipient (Figure 3B). 263 of 300 transmissions occurred within the same community, a weighted proportion of 87·1% (82·7%-91·5%). Contrary to the other variables, here there was some evidence of a difference in the proportion of transmissions coming from outside the community by trial arm (chi-squared test *p*=0·033), with 11% from outside the community in arm A, 20% in arm B and 8·3% in arm C.

### Combined characteristics of sources

We next sought to understand the extent to which characteristics of sources might occur disproportionately often in combinations. To do this, we ranked each combination according to the fraction of transmissions in which they played a role (Figure 4A). The highest fraction originated from males aged 25-40 who lived in the same community as their recipient and had no viral resistance to first-line ART. They were the sources for 1·78 times as many transmissions as the female risk group aged 20-35 with the same characteristics and 1·18 times as many transmissions as all sources outside these two age groups. There was no group that disproportionately combined several of the risk factors, as real values were generally very close to values expected if risk factors present in the population of sources had been randomly assigned to this population (Figure 4B).

**Figure 4:**
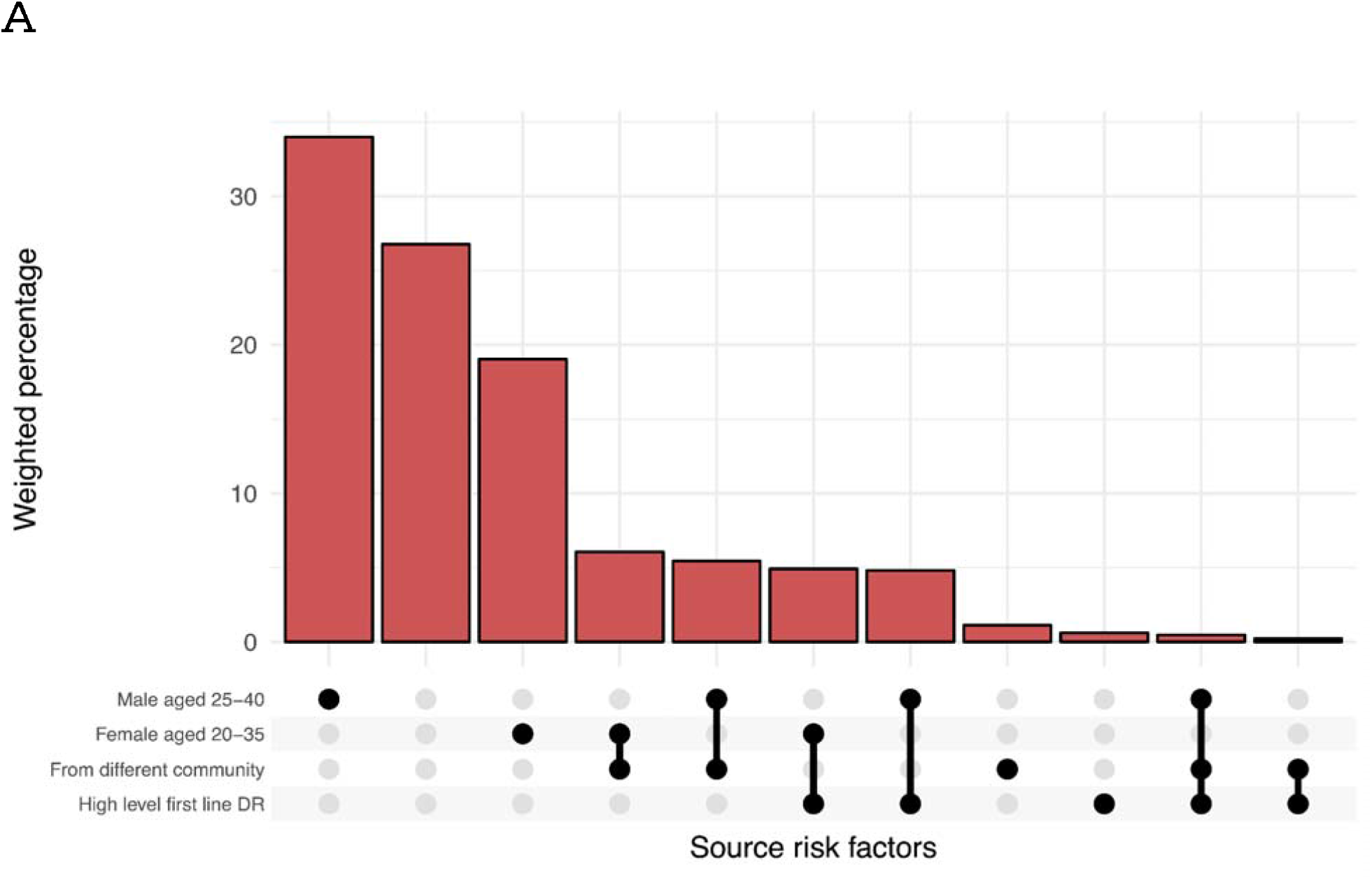
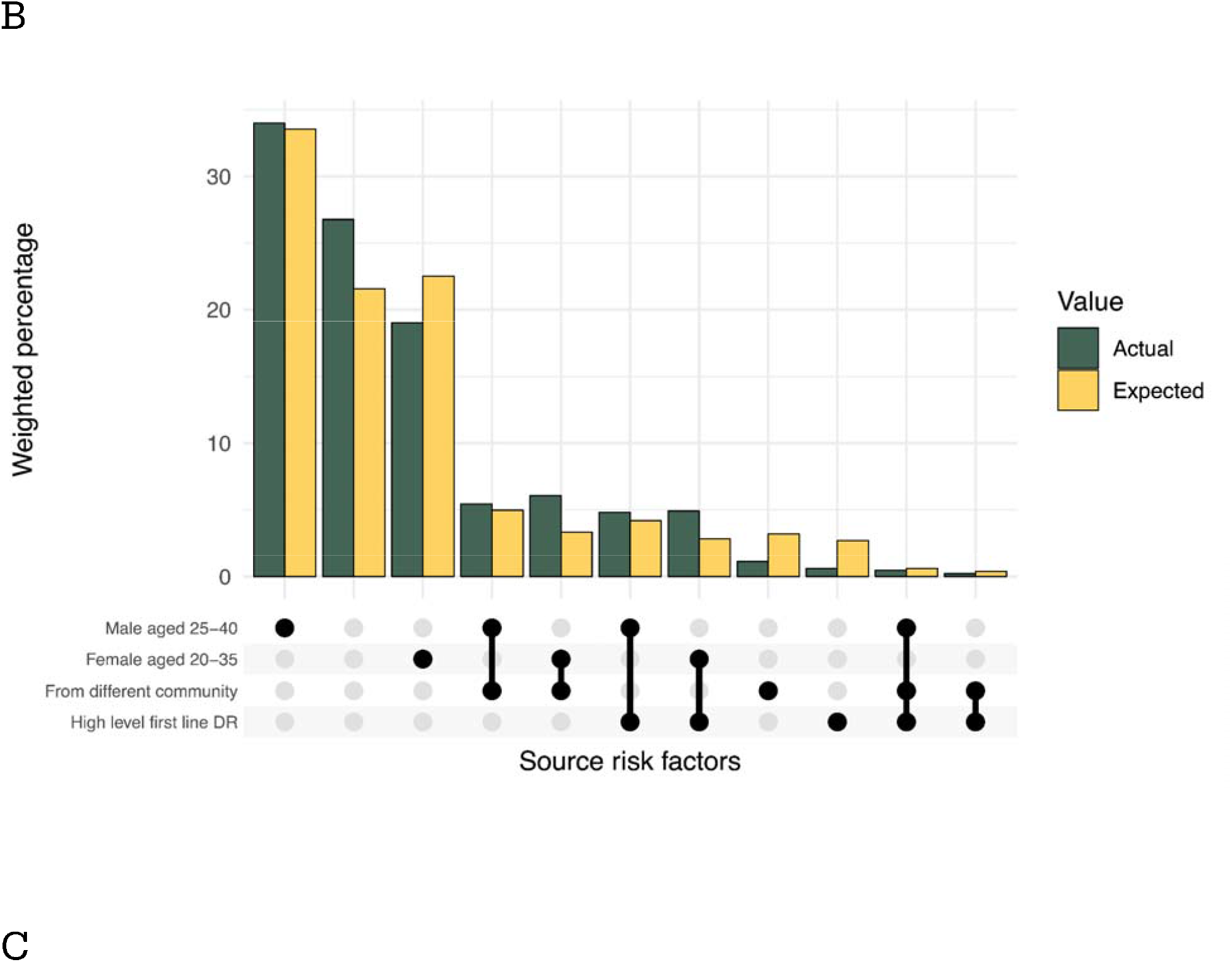
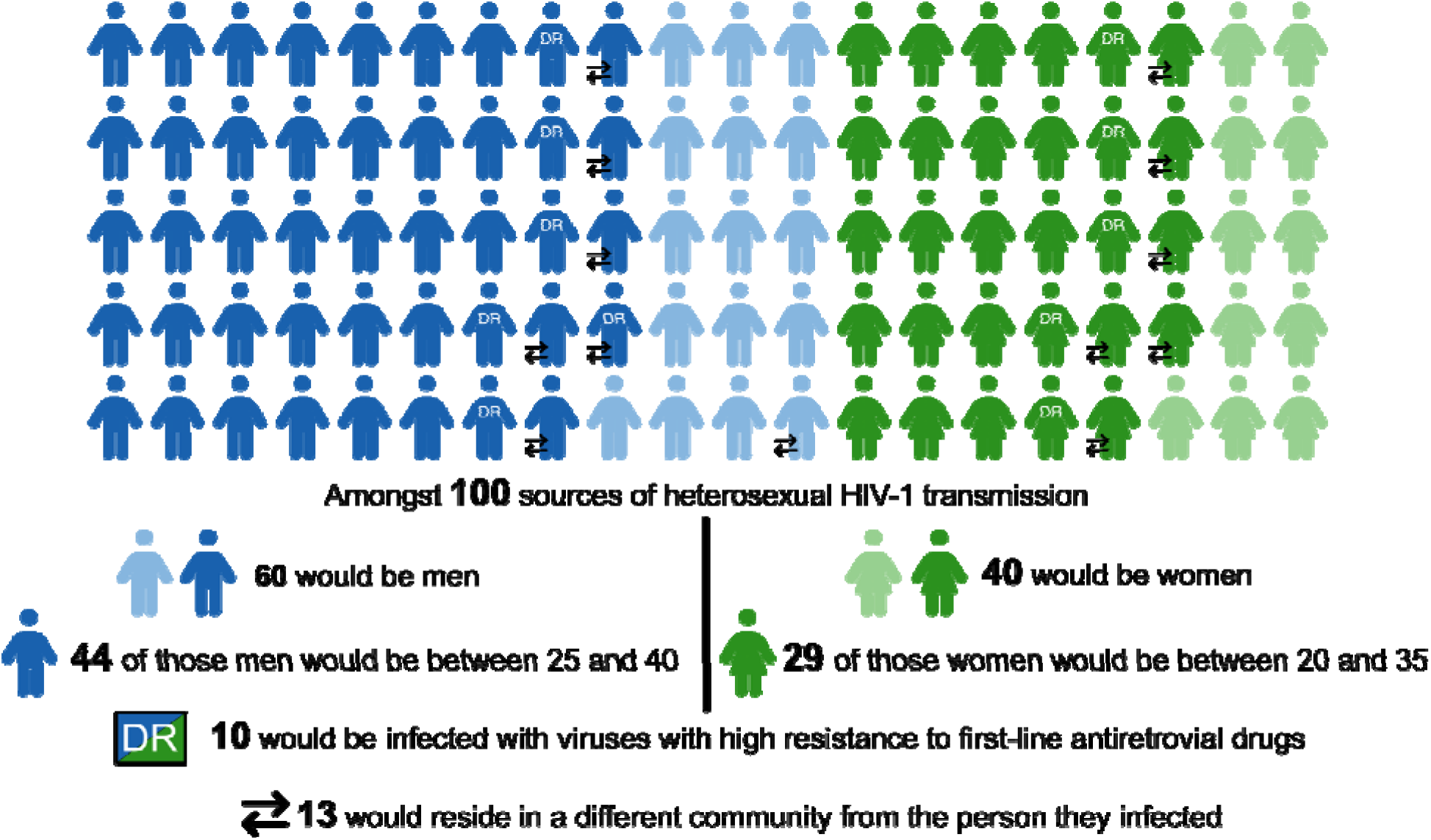
Combined characteristics of source individuals. (A) Distribution of all combinations of four key risk factors amongst the set of directed opposite-sex pairs determined to have been infected during the trial period. Each bar represents a group of sources whose characteristics are defined by the combination of black dots on the x-axis. Bars are weighted as in previous plots. (B) Actual distribution of risk factors compared to the distribution expected if they had been allocated randomly to sources based on risk factor percentages identified in the source population. (C) Pictorial representation of the expected characteristics of 100 sources of transmission. Note that as all transmission is assumed to be heterosexual, the 60 men and 40 women will have infected, respectively, 60 women and 40 men.

## Discussion

We identified characteristics of likely sources of HIV-1 transmission in Zambia between 2014 and 2019. The aim of the study is to provide policy makers with data on where transmission is still occurring in a generalised epidemic in SSA, despite delivery of a universal testing and treatment intervention, and advise where prevention efforts should be focused.

We identified men aged 25-40 as the group most commonly transmitting HIV-1. They were the source of 43·2% of transmissions (figure 4C), despite males 25-39 representing only 19·4% of prevalent cases in 2017. 30·3% of transmissions were attributable to women aged 20-35. Age distributions of sources were wide, suggesting that transmitters were not confined to any small demographic category. 2.09 times as many infections were attributable to male sources as to females, and when stratified by age this number rose to 4·23 for men and women in the 30-34 year age group and to 5·98 in the 35-39 year age group. Men in this age group were less likely than other demographic groups to be linked to care and on ART^23^. A similar phylogenetics study conducted as part of the Ya Tsie trial in Botswana largely concurred with this finding, although estimated ages for sources were somewhat greater^24^. That study, however, recorded ages at time of sampling, with no attempt to estimate time of infection. These results highlight the need for more intensive efforts in UTT programmes to achieve high and sustained ART coverage and testing for men aged 25-40, and suggest that pre-exposure prophylaxis services should also be considered for their partners^25^.

Much prevention effort on HIV in SSA has focused on intergenerational relationships, older men and the ‘sugar daddy’ phenomenon^26^. A previous phylogenetic study further suggested that the epidemic is maintained through a renewal process, in which young women are infected by considerably older men, who had been infected by women of a similar age to themselves^27^. Policies aimed at breaking this cycle have been promoted by UNAIDS and PEPFAR, in particular with the DREAMS programme. We find little evidence for this cycle or for intergenerational relationships being major drivers of transmission, with the median age of female sources being 25 and that of males 32. While we do find that a significant minority of transmissions came from outside the community (see below), this is nevertheless a minority, casting doubt on another commonly hypothesised driver of new transmissions, human mobility^28^. Our results in totality indicate that infections are predominantly local and disproportionately from men.

10·4% and 6·4% of sources’ viral populations were dominated by variants with high-level and low-level resistance against first-line ART, respectively. A caveat is that the precise timing of ART initiation was not known, and therefore some resistance could have been acquired in sources following transmission to the recipient. Nevertheless, this level of DR mirrors the high percentage of first-line ART resistance reported by other studies in the past few years^29^. Resistance against current first-line ART, including dolutegravir, needs to be monitored by local surveillance teams as the availability of HIV drug resistance testing and NGS sequencing in sub-Saharan Africa increases.

While a large majority of transmissions occurred between partners from the same community, 13% did not (figure 4C). This analysis cannot help but exclude pairs for which the source resided outside the trial population and thus could never be sampled, and so this estimate will be lower than the overall proportion of imports (and potentially much lower). Though movement does not necessarily compromise the effect of treatment as prevention, it does affect the ability of trials such as HPTN 071 to accurately measure its effect. Thus, as the primary analysis of the trial^2^ took no account of between-community transmission, even this probably underestimated figure indicates that HIV-1 incidence would have been further reduced if the PopART prevention package had been implemented country-wide. The complex statistical corrections required to assess the impact of the intervention taking into account between-community transmission are outside the scope of this study, but further genomic analysis is likely to lead to a higher estimate of its effectiveness.

A limitation of the *phyloscanner* procedure is that it is not able to rule out missing intermediaries in transmission, but for heterosexual epidemics this is easy to detect as it would result in observed same-sex pairs. Our 242 female-to-female pairs are exceptionally unlikely to represent genuine sexual transmission, but the 90 male-to-male examples could potentially be men who have sex with men (MSM). However, of all combinations of females in the dataset, a proportion of 5·18×10^−5^ (95% binomial confidence interval 4·58×10^−5^-5·87×10^−5^) were linked by the analysis, compared to 4·09×10^−5^ (3·32×10^−5^-5·03×10^−5^) of males. As the female-to-female links must be erroneous, the overlapping confidence intervals suggest no evidence for an excess of male-to-male pairs that would imply a substantial contribution of cryptic MSM transmission to infections in the sampled population. We may well underestimate the amount of MSM transmission due to simple undersampling of MSM, but this result nevertheless suggests that MSM are not widely integrated into heterosexual networks. The restriction to same-sex pairs also naturally removes any pairs with a single missing intermediary, increasing the specificity of *phyloscanner* as only situations of two or more missing intermediaries cannot be ruled out.

This study has limitations. This work was conducted in the context of the HPTN 071 trial, which potentially limits the generalisability of results to other settings. However, the trial did have a control arm, and the only variable that we explored whose distribution showed a clear difference between arms was transmission from outside the community, which was higher in intervention arms. This is consistent with the intervention reducing transmission within-community, resulting in a larger fraction of infections being caused by introductions. Nevertheless, we acknowledge that the conditions of even the control communities may have differed from those in other locations where no trial was taking place, for example because the presence of trial staff in the community might have some effect on healthcare-seeking behaviour in the population.

While sampling fractions were high, reaching between 4·9% and 9·5% of people living with HIV in the study communities, the sample was still not fully representative of the whole HIV-positive population. In particular, the design restricted sampled individuals to one person per household in the PC, limiting our ability to identify within-household transmission pairs. This is mitigated by the collection of samples in the HCFs, which had no such restrictions, but may have introduced its own bias towards identifying individuals who presented to these facilities of their own volition. The sequencing approach only yielded sequences for participants with more than 5000 viral copies/mL. This will exclude patients with lower viral loads, but still corresponds to 92·8% of participants with viral loads above 1500 copies/mL^30^. We estimate that the contribution of MSM to transmission amongst the sampled members of this community is at most small, and hence that by including only opposite-sex pairs we analysed the transmission route responsible for sustaining infections in this population. Nevertheless, it remains possible that genuine examples of MSM transmission were detected, and we did not explore the characteristics of those pairs as it was impossible to identify which they were. A proper investigation of any MSM transmission in this or similar populations would require a different study design.

HPTN 071-02 (PopART) Phylogenetics is the largest HIV phylogenetic study conducted to date, the first large transmission study to be based on an a priori power calculation, and the most comprehensive study of characteristics of sources of HIV-1 infection in SSA. The study highlights that residual transmission was not limited to coming from small risk groups. Men aged 25-40 however were, as a group, responsible for a large share of transmissions and should be prioritised in prevention efforts, even if linking them to care requires more effort.

## Data Availability

Data are available via the PANGEA-HIV consortium (www.pangea-hiv.org) upon request.

## Acknowledgements

We thank all of the field staff and everybody working with the research team in Zambia for their contribution to this study.

## Author contributions

Conceptualization: CF, DBu, HA, MC, MS, OR, RH, SE, SF

Data curation: AS, CF, DD, TG

Formal Analysis: CF, DBo, MH, TG, WP, NO

Funding acquisition: CF, DBu, HA, MC, OR, RH, SF

Investigation: DBo, GMC, MdC,

Methodology: DBo, MH, TG

Project administration: ABC, BK, CF, DBo, EPM, LAD, ML, SE, SF

Software: DBo, MH, TG

Supervision: CF, HA

Dissemination of findings: HA, MS

Visualization: MH

Writing – original draft: CF, LAD, MH

Writing – review & editing: BK, DBo, DBu, MC, ML, RH, SE, SF, TG

## Data sharing

Consensus sequences with PANGEA-specific identifiers, sex of participant and year and country of sampling will be made available on Mendeley Data upon publication and on GenBank as soon as possible. A data dictionary for these fields, the consent form and the analysis code will also be made available on Mendeley Data upon publication.

Additional data fields and/or NGS sequencing data can be requested via the data sharing procedures of the PANGEA consortium (see www.pangea-hiv.org). The requests are assessed by the PANGEA Steering Committee and data will usually be made available for non-commercial purposes unless there is a risk of participants and communities being harmed or stigmatised, or participants’ privacy being compromised. If multiple applicants are aiming to pursue the same analyses, they will be encouraged to coordinate efforts.

