## Supplement for "Demographics of people who transmit HIV-1 in Zambia: a molecular epidemiology analysis in the HPTN-071 PopART study"

**Appendix**

**Table of contents**

1. Phyloscanner procedure
2. Reconstruction of direction of transmission by estimated time of infection
3. Power calculation for transmission pairs
4. Detection and classification of drug resistance mutations (further details)
5. Calculation of relative transmission rates
6. Sensitivity analysis
7. Table S1: Extended participants table
8. Table S2: Extended baseline demographics table
9. Table S3: Sensitivity analysis results
10. Table S4: Command line options for HIV-TRACE, IQ-TREE, and *phyloscanner*
11. References

**Phyloscanner procedure**

We used a set of 898 genomic windows, each of 250 base pairs in length, spaced at regular intervals to span the entire HIV-1 genome. Alignment of the reads intersecting each window, together with a set of 22 reference sequences, was performed with *phyloscanner*, and phylogenetic reconstruction performed on each alignment using IQ-TREE 1.6.12^1^ and the FreeRate nucleotide substitution model with six rate categories. For a full list of command-line options used in HIV-TRACE, *phyloscanner* and IQ-TREE, see Table S4.

Likely transmission pairs were identified from the 898 phylogenies where their subtrees lay within a normalised patristic distance threshold of 0.02 substitutions per site in at least 50% of windows, after adjusting for windows with missing coverage as in^2^. Topological determination of the direction of transmission also used *phyloscanner*. Directionality was called if it was established in at least 33% of windows (adjusted for missing windows as before).

In a small number of cases where participants were reconstructed as the probable recipient of more than one transmission, the source was taken as the one with the greatest number of *phyloscanner* windows with normalised patristic distance less than 0.02.

**Reconstruction of direction of transmission by estimated time of infection**

Phylo-TSI gives both a point estimate for the date of infection and a standard deviation of the estimate; these were used to fit normal distributions. Directionality was called if the distributions for the two infection date estimates had an overlap of less than 20% area under the curve.

**Power calculation for transmission pairs**

The power calculation outlined in the PopART Phylogenetics study protocol (p55-57 of ^3^) predicted that a total of 269 transmission pairs from incident cases would be identifiable, under the assumption that of all the transmission pairs for which both individuals provided sequences, 75% could be identified as pairs by the analysis. In the event, we identified 300 such pairs. This is despite the dataset consisting of 6,865 sequences, considerably lower than the protocol’s prediction of the acquisition of 9,156 (p31).

**Detection and classification of drug resistance mutations (further details)**

A bioinformatic pipeline, drmSEQ, was used to predict drug resistance to first-line adult ART based on detection of mutations in the Illumina reads generated by veSEQ-HIV using the Stanford HIV Drug Resistance Database scoring system (HIVdb version 8.9.1)^4^ as follows: wild type/susceptible for scores 0-14 -, low-level resistance for scores 15-29, and high-level resistance for scores 30 and above. The method has been previously validated against an FDA-approved drug resistance assay^5^. First-line adult ART national guidance at the time of sampling was non-nucleoside inhibitor efavirenz in combination with nucleoside inhibitors including abacavir, AZT, D4T, DDI, FTC, 3TC or Tenofovir.

Mutations were reported when detected in three or more PCR-deduplicated reads and 5% or more reads spanning each site. Drug Resistance was reported as ‘unknown’ when fewer than 50% of sites relevant to each drug reached the minimum coverage threshold of 3 or more PCR-deduplicated reads.

**Calculation of relative transmission rates**

The relative contribution of demographic groups to transmission, compared to their share of the overall population, was determined as follows. Suppose
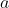
 represent an age group and
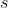
 a sex. We calculated
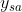
, the proportion of all HIV+ individuals in 2017 belonging to
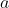
 and
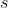
 according to the IBM, and
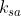
, the proportion of our pairs, weighted as described above, where the source belonged to
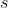
 and the recipient belonged to
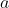
 in July 2017 (regardless of their estimated date of infection). We then calculated:

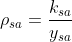

This represents the relative rate of transmissions coming from the combination of
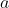
 and
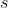
. It is above 1 if that demographic category is overrepresented amongst sources, and less than 1 if it is underrepresented.

We can also calculate
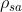
 across all age groups to get the statistics
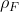
 for all females and
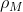
 for all males. Then

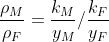

represents the ratio of the number of male sources per infected male to the number of female sources per infected female. This ratio can also be calculated individually in each age group.

For
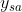
 we used the IBM-derived HIV+ population. As the vast majority of new infections will come from people not currently on ART, an alternative statistic is
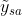
, representing the proportion of HIV positive individuals not currently on ART who are in both
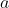
 and
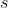
. The relative rate using
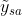
 as a denominator,
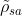
, is calculated in the same way.

To derive confidence intervals for these statistics, we performed a non-parametric bootstrap by sampling the set of observed pairs 200 times with replacement, re-weighting this new dataset by iterative proportional fitting, and calculating
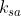
 again.

**Sensitivity analysis**

We performed a sensitivity analysis by restricting only to pairs where both source and recipient were enrolled from the HCFs. The results did not vary greatly from the main analysis (Table S3).

**Table S1. Extended participants table.** HIV-positive study participants, sequence availability, and inferred sources and recipients in probable transmission pairs, by arm of the study. HCF = health care facilities, PC seroconverters = participants in the population cohort who seroconverted during the trial period. PC0, PC12, PC24 = population cohort participants, HIV positive at baseline, recruited at the start of the trial (PC0) after 12 months (PC12N) and after 24 months (PC24N).

| Recruitment | Total HIV positive participants | Participants successfully sequenced | Sequences meeting quality threshold | Sources | Recipients |
| --- | --- | --- | --- | --- | --- |
| HCF | 5728 | 5319 | 4686 | 258 | 274 |
| PC seroconverters | 453 | 180 | 171 | 2 | 17 |
| PC0 | 3065 | 843 | 512 | 18 | 2 |
| PC12N | 738 | 247 | 110 | 3 | 2 |
| PC24N | 428 | 275 | 133 | 3 | 5 |

**Table S2. Demographic and other characteristics of all eligible participants, and sources and recipients in the reconstructed transmission pairs, by trial arm and overall.** A total of 300 pairs were found, but 14 sources had multiple probable recipients.

| **Variable** | **Value** | **All eligible participants** | | | | **Sources** | | | | **Recipients** | | | |
| --- | --- | --- | --- | --- | --- | --- | --- | --- | --- | --- | --- | --- | --- |
|  |  | **Arm** | | | **Total** | **Arm** | | | **Total** | **Arm** | | | **Total** |
|  |  | **A** | **B** | **C** |  | **A** | **B** | **C** |  | **A** | **B** | **C** |  |
| Cohort | HCF | 1474 (81.8%) | 1492 (84.9%) | 1719 (83.8%) | 4685 (83.5%) | 85 (91.4%) | 68 (85.0%) | 105 (94.6%) | 258 (90.8%) | 85 (85.9%) | 83 (96.5%) | 106 (92.2%) | 274 (91.3%) |
|  | PC | 328 (18.2%) | 266 (15.1%) | 332 (16.2%) | 926 (16.5%) | 8 (8.6%) | 12 (15.0%) | 6 (5.4%) | 26 (9.2%) | 14 (14.1%) | 3 (3.5%) | 9 (7.8%) | 26 (8.7%) |
| Sex | F | 1100 (61.0%) | 1033 (58.8%) | 1201 (58.6%) | 3334 (59.4%) | 43 (46.2%) | 38 (47.5%) | 40 (36.0%) | 121 (42.6%) | 53 (53.5%) | 47 (54.7%) | 70 (60.9%) | 170 (56.7%) |
|  | M | 702 (39.0%) | 725 (41.2%) | 850 (41.4%) | 2277 (40.6%) | 50 (53.8%) | 42 (52.5%) | 71 (64.0%) | 163 (57.4%) | 46 (46.5%) | 39 (45.3%) | 45 (39.1%) | 130 (43.3%) |
| Year of birth | 1935-1939 | 0 (0.0%) | 1 (0.1%) | 2 (0.1%) | 3 (0.1%) | 0 (0.0%) | 0 (0.0%) | 0 (0.0%) | 0 (0.0%) | 0 (0.0%) | 0 (0.0%) | 1 (0.9%) | 1 (0.3%) |
|  | 1940-1944 | 1 (0.1%) | 3 (0.2%) | 1 (0.0%) | 5 (0.1%) | 0 (0.0%) | 0 (0.0%) | 0 (0.0%) | 0 (0.0%) | 0 (0.0%) | 0 (0.0%) | 0 (0.0%) | 0 (0.0%) |
|  | 1945-1949 | 5 (0.3%) | 3 (0.2%) | 6 (0.3%) | 14 (0.2%) | 0 (0.0%) | 0 (0.0%) | 0 (0.0%) | 0 (0.0%) | 0 (0.0%) | 0 (0.0%) | 0 (0.0%) | 0 (0.0%) |
|  | 1950-1954 | 13 (0.7%) | 7 (0.4%) | 8 (0.4%) | 28 (0.5%) | 1 (1.1%) | 1 (1.2%) | 0 (0.0%) | 2 (0.7%) | 0 (0.0%) | 0 (0.0%) | 0 (0.0%) | 0 (0.0%) |
|  | 1955-1959 | 23 (1.3%) | 28 (1.6%) | 22 (1.1%) | 73 (1.3%) | 0 (0.0%) | 1 (1.2%) | 0 (0.0%) | 1 (0.4%) | 2 (2.0%) | 2 (2.3%) | 0 (0.0%) | 4 (1.3%) |
|  | 1960-1964 | 34 (1.9%) | 43 (2.4%) | 45 (2.2%) | 122 (2.2%) | 1 (1.1%) | 1 (1.2%) | 1 (0.9%) | 3 (1.1%) | 1 (1.0%) | 2 (2.3%) | 0 (0.0%) | 3 (1.0%) |
|  | 1965-1969 | 71 (3.9%) | 70 (4.0%) | 80 (3.9%) | 221 (3.9%) | 5 (5.4%) | 1 (1.2%) | 4 (3.6%) | 10 (3.5%) | 2 (2.0%) | 5 (5.8%) | 2 (1.7%) | 9 (3.0%) |
|  | 1970-1974 | 137 (7.6%) | 149 (8.5%) | 175 (8.5%) | 461 (8.2%) | 10 (10.8%) | 4 (5.0%) | 6 (5.4%) | 20 (7.0%) | 3 (3.0%) | 1 (1.2%) | 3 (2.6%) | 7 (2.3%) |
|  | 1975-1979 | 241 (13.4%) | 242 (13.8%) | 288 (14.0%) | 771 (13.7%) | 7 (7.5%) | 8 (10.0%) | 18 (16.2%) | 33 (11.6%) | 14 (14.1%) | 11 (12.8%) | 6 (5.2%) | 31 (10.3%) |
|  | 1980-1984 | 359 (19.9%) | 353 (20.1%) | 405 (19.7%) | 1117 (19.9%) | 18 (19.4%) | 23 (28.7%) | 26 (23.4%) | 67 (23.6%) | 11 (11.1%) | 14 (16.3%) | 23 (20.0%) | 48 (16.0%) |
|  | 1985-1989 | 370 (20.5%) | 358 (20.4%) | 437 (21.3%) | 1165 (20.8%) | 21 (22.6%) | 15 (18.8%) | 21 (18.9%) | 57 (20.1%) | 25 (25.3%) | 15 (17.4%) | 33 (28.7%) | 73 (24.3%) |
|  | 1990-1994 | 365 (20.3%) | 348 (19.8%) | 391 (19.1%) | 1104 (19.7%) | 21 (22.6%) | 21 (26.2%) | 22 (19.8%) | 64 (22.5%) | 35 (35.4%) | 26 (30.2%) | 29 (25.2%) | 90 (30.0%) |
|  | 1995-1999 | 180 (10.0%) | 152 (8.6%) | 186 (9.1%) | 518 (9.2%) | 9 (9.7%) | 5 (6.2%) | 13 (11.7%) | 27 (9.5%) | 6 (6.1%) | 10 (11.6%) | 18 (15.7%) | 34 (11.3%) |
|  | 2000-2004 | 2 (0.1%) | 0 (0.0%) | 0 (0.0%) | 2 (0.0%) | 0 (0.0%) | 0 (0.0%) | 0 (0.0%) | 0 (0.0%) | 0 (0.0%) | 0 (0.0%) | 0 (0.0%) | 0 (0.0%) |
|  | Unknown | 1 (0.1%) | 1 (0.1%) | 5 (0.2%) | 7 (0.1%) | 0 (0%) | 0 (0%) | 0 (0%) | 0 (0%) | 0 (0%) | 0 (0%) | 0 (0%) | 0 (0%) |
| Year of sampling | 2013 | 1 (0.1%) | 0 (0.0%) | 0 (0.0%) | 1 (0.0%) | 0  (0%) | 0  (0%) | 0  (0%) | 0  (0%) | 0  (0%) | 0  (0%) | 0  (0%) | 0  (0%) |
|  | 2014 | 124 (6.9%) | 148 (8.4%) | 147 (7.2%) | 419 (7.5%) | 4 (4.3%) | 9 (11.2%) | 3 (2.7%) | 16 (5.6%) | 0 (0.0%) | 0 (0.0%) | 1 (0.9%) | 1 (0.3%) |
|  | 2015 | 48 (2.7%) | 44 (2.5%) | 52 (2.5%) | 144 (2.6%) | 2 (2.2%) | 0 (0.0%) | 1 (0.9%) | 3 (1.1%) | 3 (3.0%) | 0 (0.0%) | 2 (1.7%) | 5 (1.7%) |
|  | 2016 | 466 (25.9%) | 651 (37.0%) | 586 (28.6%) | 1703 (30.4%) | 21 (22.6%) | 30 (37.5%) | 33 (29.7%) | 84 (29.6%) | 29 (29.3%) | 36 (41.9%) | 27 (23.5%) | 92 (30.7%) |
|  | 2017 | 850 (47.2%) | 717 (40.8%) | 916 (44.7%) | 2483 (44.3%) | 48 (51.6%) | 34 (42.5%) | 48 (43.2%) | 130 (45.8%) | 50 (50.5%) | 40 (46.5%) | 53 (46.1%) | 143 (47.7%) |
|  | 2018 | 313 (17.4%) | 198 (11.3%) | 350 (17.1%) | 861 (15.3%) | 18 (19.4%) | 7 (8.8%) | 26 (23.4%) | 51 (18.0%) | 17 (17.2%) | 10 (11.6%) | 32 (27.8%) | 59 (19.7%) |
| Marital status  at sampling | Never married | 314 (17.4%) | 277 (15.8%) | 382 (18.6%) | 973 (17.3%) | 12 (12.9%) | 8 (10.0%) | 13 (11.7%) | 33 (11.6%) | 10 (10.1%) | 14 (16.3%) | 16 (13.9%) | 40 (13.3%) |
|  | Married/living as married | 970 (53.8%) | 975 (55.5%) | 1141 (55.6%) | 3086 (55.0%) | 66 (71.0%) | 60 (75.0%) | 83 (74.8%) | 209 (73.6%) | 68 (68.7%) | 56 (65.1%) | 80 (69.6%) | 204 (68.0%) |
|  | Divorced/  separated | 360 (20.0%) | 373 (21.2%) | 371 (18.1%) | 1104 (19.7%) | 14 (15.1%) | 11 (13.8%) | 12 (10.8%) | 37 (13.0%) | 16 (16.2%) | 14 (16.3%) | 14 (12.2%) | 44 (14.7%) |
|  | Widowed | 147 (8.2%) | 125 (7.1%) | 145 (7.1%) | 417 (7.4%) | 1 (1.1%) | 1 (1.2%) | 1 (0.9%) | 3 (1.1%) | 5 (5.1%) | 2 (2.3%) | 5 (4.3%) | 12 (4.0%) |
|  | Unknown | 11 (0.6%) | 8 (0.5%) | 12 (0.6%) | 31 (0.6%) | 0 (0.0%) | 0 (0.0%) | 2 (1.8%) | 2 (0.7%) | 0 (0%) | 0 (0%) | 0 (0%) | 0 (0%) |
| Estimated elapsed years  from infection to sampling | 0 | 362 (20.1%) | 280 (15.9%) | 344 (16.8%) | 986 (17.6%) | 11 (11.8%) | 9 (11.2%) | 16 (14.4%) | 36 (12.7%) | 54 (54.5%) | 41 (47.7%) | 56 (48.7%) | 151 (50.3%) |
|  | 1 | 289 (16.0%) | 283 (16.1%) | 309 (15.1%) | 881 (15.7%) | 12 (12.9%) | 11 (13.8%) | 15 (13.5%) | 38 (13.4%) | 32 (32.3%) | 24 (27.9%) | 39 (33.9%) | 95 (31.7%) |
|  | 2 | 259 (14.4%) | 261 (14.8%) | 274 (13.4%) | 794 (14.2%) | 18 (19.4%) | 12 (15.0%) | 17 (15.3%) | 47 (16.5%) | 9 (9.1%) | 16 (18.6%) | 16 (13.9%) | 41 (13.7%) |
|  | 3 | 200 (11.1%) | 201 (11.4%) | 219 (10.7%) | 620 (11.0%) | 17 (18.3%) | 12 (15.0%) | 16 (14.4%) | 45 (15.8%) | 4 (4.0%) | 5 (5.8%) | 3 (2.6%) | 12 (4.0%) |
|  | 4 | 136 (7.5%) | 128 (7.3%) | 200 (9.8%) | 464 (8.3%) | 4 (4.3%) | 7 (8.8%) | 9 (8.1%) | 20 (7.0%) | 0 (0.0%) | 0 (0.0%) | 1 (0.9%) | 1 (0.3%) |
|  | 5 | 127 (7.0%) | 145 (8.2%) | 153 (7.5%) | 425 (7.6%) | 5 (5.4%) | 5 (6.2%) | 8 (7.2%) | 18 (6.3%) | 0  (0%) | 0  (0%) | 0  (0%) | 0  (0%) |
|  | 6 | 58 (3.2%) | 67 (3.8%) | 61 (3.0%) | 186 (3.3%) | 6 (6.5%) | 6 (7.5%) | 5 (4.5%) | 17 (6.0%) | 0  (0%) | 0  (0%) | 0  (0%) | 0  (0%) |
|  | 7 | 65 (3.6%) | 53 (3.0%) | 81 (3.9%) | 199 (3.5%) | 6 (6.5%) | 3 (3.8%) | 4 (3.6%) | 13 (4.6%) | 0  (0%) | 0  (0%) | 0  (0%) | 0  (0%) |
|  | 8 | 100 (5.5%) | 116 (6.6%) | 129 (6.3%) | 345 (6.1%) | 6 (6.5%) | 4 (5.0%) | 4 (3.6%) | 14 (4.9%) | 0  (0%) | 0  (0%) | 0  (0%) | 0  (0%) |
|  | 9 | 107 (5.9%) | 117 (6.7%) | 134 (6.5%) | 358 (6.4%) | 5 (5.4%) | 7 (8.8%) | 5 (4.5%) | 17 (6.0%) | 0  (0%) | 0  (0%) | 0  (0%) | 0  (0%) |
|  | 10 | 72 (4.0%) | 82 (4.7%) | 111 (5.4%) | 265 (4.7%) | 0 (0.0%) | 4 (5.0%) | 8 (7.2%) | 12 (4.2%) | 0  (0%) | 0  (0%) | 0  (0%) | 0  (0%) |
|  | 11 | 23 (1.3%) | 22 (1.3%) | 32 (1.6%) | 77 (1.4%) | 3 (3.2%) | 0 (0.0%) | 4 (3.6%) | 7 (2.5%) | 0  (0%) | 0  (0%) | 0  (0%) | 0  (0%) |
|  | 12 | 4 (0.2%) | 3 (0.2%) | 4 (0.2%) | 11 (0.2%) | 0 (0%) | 0 (0%) | 0 (0%) | 0 (0%) | 0  (0%) | 0  (0%) | 0  (0%) | 0  (0%) |

**Table S3. Sensitivity analysis.** Key quantities from the main analysis and from the two sensitivity analyses. Apart from the first row, all quantities are demographically weighted. Ranges in brackets are 95% confidence intervals.

| Quantity | Dataset | |
| --- | --- | --- |
|  | Main analysis | HCF only |
| Number of pairs | 300 | 248 |
| Median age of male sources | 32 | 32.3 |
| Median age of female sources | 24.9 | 25.4 |
| % transmissions from males aged 25-40 | 43.2 (36.8-49.7) | 47.3 (39.8-54.8) |
| % transmissions from females aged 20-35 | 29 (23.1-34.9) | 26.7 (20-33.3) |
| % transmissions from sources without DRMs | 78.8 (73.5-84.1) | 78.8 (72.7-84.9) |
| % transmissions from same community as recipient | 87.1 (82.7-91.5) | 88.4 (83.5-93.2) |

**Table S4: Command line options for HIV-TRACE, IQ-TREE, and the two phyloscanner commands.** For further details, see the relevant package manuals.

| Option | Meaning | Value | Value meaning/notes |
| --- | --- | --- | --- |
| HIV-TRACE | | | |
| --ambiguities | Handling of ambiguous nucleotides | average | Average all possible resolutions |
| --minoverlap | Minimum proportional overlap of sequences for inclusion in distance calculations | 0.5 |  |
| --threshold | Distance threshold | 0.04 |  |
| --curate | Contaminant screening | remove | Remove sequences that cluster with the reference |
| phyloscanner_make_trees | | | |
| --pairwise-align-to | Pairwise align all reads to this reference | <HXB2 sequence file> |  |
| --excision-coords | Coordinates in the reference to be excised | 823, 824, 825, 892, 893, 894, 907, 908, 909, 1012, 1013, 1014, 1156, 1157, 1158, 1384, 1385, 1386, 1444, 1445, 1446, 1930, 1931, 1932, 1957, 1958, 1959, 2014, 2015, 2016, 2023, 2024, 2025, 2080, 2081, 2082, 2134, 2135, 2136, 2191, 2192, 2193, 2280, 2281, 2282, 2283, 2284, 2285, 2298, 2299, 2300, 2310, 2311, 2312, 2316, 2317, 2318, 2319, 2320, 2321, 2322, 2323, 2324, 2340, 2341, 2342, 2346, 2347, 2348, 2349, 2350, 2351, 2352, 2353, 2354, 2355, 2356, 2357, 2358, 2359, 2360, 2373, 2374, 2375, 2379, 2380, 2381, 2385, 2386, 2387, 2388, 2389, 2390, 2391, 2392, 2393, 2394, 2395, 2396, 2400, 2401, 2402, 2409, 2410, 2411, 2412, 2413, 2414, 2415, 2416, 2417, 2424, 2425, 2426, 2430, 2431, 2432, 2436, 2437, 2438, 2439, 2440, 2441, 2442, 2443, 2444, 2457, 2458, 2459, 2460, 2461, 2462, 2463, 2464, 2465, 2469, 2470, 2471, 2472, 2473, 2474, 2478, 2479, 2480, 2481, 2482, 2483, 2496, 2497, 2498, 2499, 2500, 2501, 2502, 2503, 2504, 2505, 2506, 2507, 2514, 2515, 2516, 2517, 2518, 2519, 2520, 2521, 2522, 2526, 2527, 2528, 2529, 2530, 2531, 2535, 2536, 2537, 2670, 2671, 2672, 2679, 2680, 2681, 2703, 2704, 2705, 2709, 2710, 2711, 2733, 2734, 2735, 2742, 2743, 2744, 2748, 2749, 2750, 2751, 2752, 2753, 2754, 2755, 2756, 2757, 2758, 2759, 2769, 2770, 2771, 2772, 2773, 2774, 2778, 2779, 2780, 2811, 2812, 2813, 2814, 2815, 2816, 2817, 2818, 2819, 2823, 2824, 2825, 2841, 2842, 2843, 2847, 2848, 2849, 2850, 2851, 2852, 2856, 2857, 2858, 2865, 2866, 2867, 2871, 2872, 2873, 2892, 2893, 2894, 2895, 2896, 2897, 2901, 2902, 2903, 2904, 2905, 2906, 2952, 2953, 2954, 2961, 2962, 2963, 3000, 3001, 3002, 3015, 3016, 3017, 3018, 3019, 3020, 3030, 3031, 3032, 3042, 3043, 3044, 3084, 3085, 3086, 3090, 3091, 3092, 3099, 3100, 3101, 3111, 3112, 3113, 3117, 3118, 3119, 3135, 3136, 3137, 3171, 3172, 3173, 3177, 3178, 3179, 3180, 3181, 3182, 3189, 3190, 3191, 3192, 3193, 3194, 3204, 3205, 3206, 3210, 3211, 3212, 3222, 3223, 3224, 3228, 3229, 3230, 3237, 3238, 3239, 3246, 3247, 3248, 3249, 3250, 3251, 3255, 3256, 3257, 3261, 3262, 3263, 3396, 3397, 3398, 3501, 3502, 3503, 3546, 3547, 3548, 3705, 3706, 3707, 4425, 4426, 4427, 4449, 4450, 4451, 4503, 4504, 4505, 4518, 4519, 4520, 4590, 4591, 4592, 4641, 4642, 4643, 4647, 4648, 4649, 4656, 4657, 4658, 4668, 4669, 4670, 4671, 4672, 4673, 4692, 4693, 4694, 4722, 4723, 4724, 4782, 4783, 4784, 4974, 4975, 4976, 5016, 5017, 5018, 5067, 5068, 5069, 7863, 7864, 7865, 7866, 7867, 7868, 7869, 7870, 7871, 7872, 7873, 7874, 7875, 7876, 7877, 7881, 7882, 7883, 7884, 7885, 7886 | Known drug resistance sites in HXB2 |
| --min-read-count | Minimum count for each unique read | 1 |  |
| IQ-TREE | | | |
| -m | Substitution model | GTR+F+R6 | General Time Reversible rate model, Empirical state frequencies, FreeRate site variation model with six categories |
| phyloscanner_analyse_trees.R | | | |
| splitsRule | Parsimony reconstruction settings | s,15 | Sankoff algorithm with *k*=15 |
| --outgroupName | Outgroup name | <HXB2 sequence name> |  |
| --multifurcationThreshold | Multifurcation collapse threshold | 1E-5 |  |
| --normRefFileName | Per-window branch length normalisation file |  | Derived from a phylogeny of global HIV diversity constructed using the FreeRate model |
| --normStandardiseGagPol | Standardise normalising constants so that the average on *gag*+*pol* equals 1 |  |  |
| --parsimonyBlacklistK | Value of *k* in the parsimony algorithm when used to identify contaminant reads | 15 |  |
| --rawBlacklistThreshold | Threshold of reads below which all tips in a subgraph from a single individual will be blacklisted | 10 |  |
| --ratioBlacklistThreshold | Threshold for the proportion of all reads from a single individual that are members of a single subgraph, below which all tips from that subgraph will be blacklisted | 0.05 |  |
| --readCountsMatterOnZeroLengthBranches | Take into account read counts when applying parsimony algorithm |  |  |
| --distanceThreshold | Distance threshold, normalised by genome position, below which individuals will be identified as a likely transmission pair in a window | 0.02 |  |
| --allowMultiTrans | Allow the “multiTrans” categorisation to indicate direction of transmission |  |  |
